# The Clinical Characteristics and mortality outcomes of Atrial fibrillation complicating Heart failure with reduced ejection fraction: A prospective study from South Africa

**DOI:** 10.64898/2026.06.10.26355424

**Authors:** Nonkanyiso Mboweni, Muzi Maseko, Nqoba Tsabedze, Samantha Nel, Marketa Toman, Brenda Kagodora

## Abstract

**Background:** A growing burden of cardiovascular risk factors has raised cardiovascular disease-related mortality in Sub-Saharan Africa (SSA), driving higher prevalence of heart failure with reduced ejection fraction (HFrEF) and its complication with atrial fibrillation (AF). No prospective study has examined AF’s clinical impact on HFrEF in SSA.

**Aim:** To determine AF prevalence in HFrEF, describe HFrEF-AF clinical characteristics, and determine AF’s impact on mortality.

**Methods:** In this prospective observational study at a tertiary hospital in Johannesburg, 136 HFrEF patients were enrolled and categorised as HFrEF- SR (sinus rhythm) or HFrEF-AF. Baseline clinical characteristics and biochemistry were recorded. Comprehensive echocardiography including left atrial strain by 2D speckle-tracking was performed. Median follow-up was 30.6 months.

**Results:** AF was present in 28 patients (21%). The mean age was 58.7 ± 14.9 years (52.9% male) and differed between groups (p < 0.001). Hypertensive heart disease was the leading cause of HFrEF (36%). Compared with SR, HFrEF-AF patients had poorer health status (KCCQ -12 [16–43] vs 45 [32–60], p < 0.001) and lower left atrial strain (26.2 ± 11.3%, p < 0.001). Guideline-directed medical therapy was suboptimal in the AF group: anticoagulation use was higher than SR (60% vs 9.5%, p < 0.001) but overall inadequate; HFrEF–AF patients received lower median doses of carvedilol (15.6 mg vs 25 mg, p = 0.002) and enalapril (10 mg vs 20 mg, p = 0.004), and fewer received spironolactone (50% vs 75.3%, p = 0.013). Survival was significantly lower in HFrEF-AF (0.41 [0.22–0.61]) versus SR (0.73 [0.61–0.82], p < 0.001). Independent predictors of mortality included prior stroke, lower TAPSE and KCCQ, and higher E/e’ and heart rate.

**Conclusion:** AF is common among HFrEF patients in this SSA cohort (though lower than in high-income countries) and associates with worse clinical status, suboptimal therapy, and higher mortality.

## Introduction

As the global life expectancy increases, the incidence of heart failure and atrial fibrillation (AF) has increased and is projected to rise further (1). Heart failure (HF) is estimated to account for approximately 25.6-30% of hospital admissions cardiology units in Africa (2). Owing to common cardiometabolic risk factors such as hypertension, diabetes and coronary artery disease as well as electrophysiological and neurohormonal changes, heart failure with reduced ejection fraction (HFrEF) and AF frequently coexist and the presence of one precipitates the severity of the other. In the setting of HFrEF, AF confers a significant additional risk of morbidity and mortality compared to patients in sinus rhythm (SR) (3).

Despite a well-established synergistic link between HFrEF and AF and associated adverse outcomes, there is a paucity of original research focusing specifically on HFrEF-AF in Sub-Saharan Africa (SSA) and even fewer comparative studies comparing HFrEF-AF, and HFrEF-SR. Amongst available studies conducted in SSA, the majority focus on heterogeneous heart failure populations. Studies done in high-income countries (HIC) and SSA have identified disparities between HF and AF in the West and SSA (4–7). Therefore, this study aimed to estimate the prevalence of HFrEF-AF and evaluate the clinical impact of AF on HFrEF and mortality associated with this patient population in Johannesburg, South Africa.

## Methods

### Study design and participants

A prospective observational study was conducted at Charlotte Maxeke Johannesburg Academic Hospital (CMJAH), South Africa. Two hundred and forty-eight (248) patients were consecutively screened from a specialised out-patient heart failure clinic and in-patient cardiac admission wards between June 2019 and December 2021.

### Inclusion criteria

Patients with known HF, chronically stable and/or acutely decompensated as well as patients with new-onset (“de novo”) acute HF were enrolled. The diagnosis was based on the 2016 European Society of Cardiology Heart failure Guidelines (8). Individuals with a left ventricular ejection fraction (LVEF) of less than or equal to 45% on echocardiography were enrolled into the study. A LVEF of ≤ 45% was selected due to the subjectivity associated with LVEF measurements and the absence of evidence-based therapies for managing patients with mildly reduced ejection fraction (HFmrEF) at the study’s inception. At study enrolment, patients were screened for AF with a standard 12-lead resting electrocardiogram (ECG) and corroborated with clinical findings (9).

Patients with an LVEF greater than 45%, significant valvular heart disease, pacemaker rhythm or any other rhythm were excluded from the study

Patients were separated into two groups according to rhythm status; those with HFrEF in sinus rhythm (HFrEF-SR) and those with HFrEF and concomitant atrial fibrillation (HFrEF-AF) (Figure 1).

**Figure 1.**
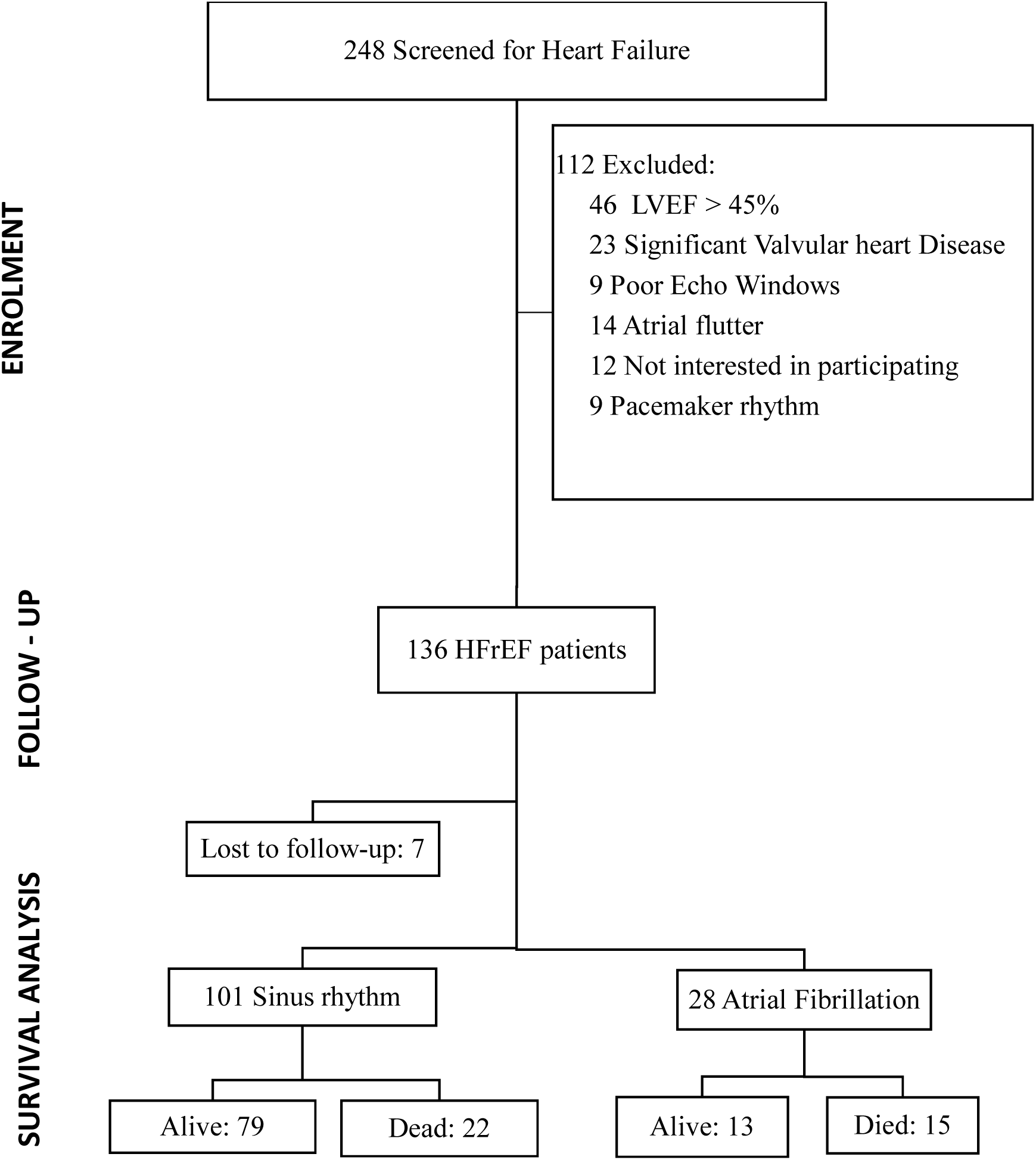
Flow diagram illustrating patient enrolment, exclusion criteria, rhythm classification, and survival status in the study cohort. A total of 248 patients were screened for heart failure. Of these, 112 were excluded due to a left ventricular ejection fraction (LVEF) > 45%, significant valvular heart disease, poor echocardiographic windows, atrial flutter, pacemaker rhythm, or lack of consent. The final cohort included 136 patients with heart failure with reduced ejection fraction (HFrEF). During follow-up, 7 patients were lost. Of the remaining 129 patients, 101 were in sinus rhythm (SR) and 28 had atrial fibrillation (AF). Survival analysis showed that 22 patients in SR and 15 in AF had died by the end of the study period.

### Data Collection

A comprehensive history and clinical examination was carried out on all patients. Heart failure symptom severity was assessed using the New York Heart Association (NYHA) functional class, and the Kansas City Cardiomyopathy Questionnaire (KCCQ) -12 was administered to evaluate the quality of life within the preceding 2 weeks of presentation (10). The Congestive heart failure, Hypertension, Age ≥75 years, Diabetes mellitus, Stroke, Vascular disease, Age. 65-74 years, sex category (female) score (CHA_2_DS_2_-VASc) was determined to assess stroke risk in patients with AF (11). Participants were subjected to clinical history taking and a physical examination focused on identifying heart failure signs and symptoms. Clinical investigations on all patients included a 12-lead ECG and a 2D transthoracic echocardiogram with doppler and speckle tracking.

### Etiological diagnosis

Treating physicians and cardiologist confirmed HF etiology using available clinical, laboratory and echocardiographic data. A diagnosis of hypertensive heart disease was based on the presence of echocardiogram abnormalities such as left ventricular hypertrophy (concentric or eccentric), increased left ventricular mass index, enlarged left ventricular or left atrial (LA) size and increased volumes and diastolic or systolic left ventricular dysfunction in patients with hypertension. The diagnosis of ischemic heart disease was made on the basis of a confirmed diagnosis of coronary artery disease, which was confirmed on history or diagnostic coronary angiogram. All patients with HFrEF at CMJAH underwent diagnostic coronary angiograms as part of a routine etiological work-up. Alcohol induced cardiomyopathy diagnosis was considered where an excessive intake (80 to 90 standard units per day) for a minimum of 5 years was documented (12, 13). Idiopathic dilated cardiomyopathy was considered in the presence of left ventricular dilatation and left ventricular systolic dysfunction in the absence hypertension, valvular heart disease, (abnormal loading conditions), coronary artery disease or any obvious clinical condition sufficient to cause global systolic impairment (14). In this cohort, we were unable to determine familial dilated cardiomyopathy diagnosis.

### Biomarker analysis and Biochemistry

Two venous blood samples (3 mL each) were collected from each patient. Samples were centrifuged and plasma collected and stored for subsequent biomarker analysis. Plasma concentrations of the N-terminal pro B type naturetic peptide (NT-proBNP) and cardiac troponin T (cTnT) were analysed using commercial sandwich enzyme-linked immunosorbent assay (ELISA) kits with a colorimetric detection,(Elabscience,Wuhan, Hubei, China) following the manufacturer’s instructions.

Baseline biochemical parameters (haemoglobin, white cell count, creatinine, potassium, sodium and lipid lipogram) were obtained from records within the last week of presentation in the event these records were available. The Modification of Diet in Renal Disease (175 × plasma creatinine−1.154 × age−0.203 [× 0.742 if female; × 1.21 if black]) calculation was used to calculate the estimated glomerular filtration rate (15).

#### Echocardiography

A detailed two dimensional transthoracic echocardiographic assessment was performed using a Philips CX50 ultrasound system (Eindhoven, Netherlands). Left ventricular systolic and diastolic function, right ventricular function and valvular structure and function were evaluated in accordance with the American Society of Echocardiography guidelines (16). Left ventricular ejection fraction was assessed using the modified biplane Simpson’s method using the apical two-chamber and four-chamber views.

##### Left atrial strain

Left atrial strain was measured using two-dimensional speckle tracking echocardiography (2D-STE) (17). Left atrial speckle tracking necessitated a minimum of three consecutive heart cycles using a frame rate of 50-80 frames per second. The endocardial border of the left atrium (LA) was traced manually, in the apical 4-chamber and 2-chamber views. The starting point for LA strain analysis was set at the onset of the QRS-complex. Reservoir strain was defined as the peak positive strain value, contraction strain was defined as the peak value at the onset of the p-wave from the ECG trace, and conduit strain was calculated as the difference between reservoir and contraction strain. In patients with AF, the conduit and contraction strain were not measured due to the absence of atrial pump function (18).

Images were acquired, stored and analysed offline using a commercially available Philips QLAB 13 (Eindhoven, Netherlands) software.

#### Follow-up and clinical outcome data

Participants were followed up telephonically, on a three-monthly basis until June 2024. The outcome of interest was all-cause mortality.

#### Statistical analysis

Demographic, clinical and biochemical characteristics are presented in frequency tables and stratified according to the presence and absence of AF. Shapiro-Wilk test was used to determine the distribution among continuous variables. Normally distributed data were reported as mean ±standard deviation and median (interquartile range) for non-parametric data and number (percentages) for categorical data. Categorical data were compared using Pearson’s chi-square test or Fisher’s exact test. For normally distributed continuous variables, a student t-test was performed and a Wilcoxon rank sum test was employed for non-parametric data. For all-cause mortality, a univariate logistic regression model was used to calculate the unadjusted odds ratio for each variable of interest, and a p-value of <0.05 qualified as significant to use these variables to build a multivariate logistic regression model. Multicollinearity diagnostics were conducted and variables with a variance inflation factor (VIF) greater than five were removed from the model (19). Backwards stepwise logistic regression was employed to compute the final model. Overall survival was measured from the date of enrolment to the date of death or the last follow-up date in June 2024. Times for patients without the event of interest were censored at the last date the patient was known to be alive. Survival curves were plotted by the Kaplan-Meier method. A log-rank test was used to compare survival probability between AF and SR patients. A statistical significance threshold of 5% and a confidence interval of 95% was used. Statistical analyses were performed using STATA (version 17, College Station, Texas).

#### Ethics declaration

Approval to conduct the study was granted by the University of the Witwatersrand Human Research Ethics Committee (Ethics Clearance certificate number: M190349). The study complied with the Declaration of Helsinki, and written informed consent was obtained from all study patients.

## Results

### Demographic and clinical characteristics

A total of 248 patients were screened in the current study. One hundred and eleven patients were excluded, leaving 136 patients included in the final analysis (Figure. 1). Patients lost to follow-up were excluded from the survival analysis. The baseline demographic and clinical characteristics are summarised in Tables I. Males represented a majority (52.9%) of participants and the mean age of participants was 58.7±14.9 years. One hundred (74%) of patients were of Black African ethnicity (p< 0.001) and a greater percentage of Caucasians (35%) were present in the HFrEF-AF group compared to the HFrEF-SR group (8.3%) (Table I). Hypertensive heart disease was the leading cause of HFrEF (33.8%) followed by idiopathic dilated cardiomyopathy (19.1%) and ischemic heart disease (19.1%). The median duration of HFrEF was 3.6 [1.1-7.1] years (Table I).

**Table I.**
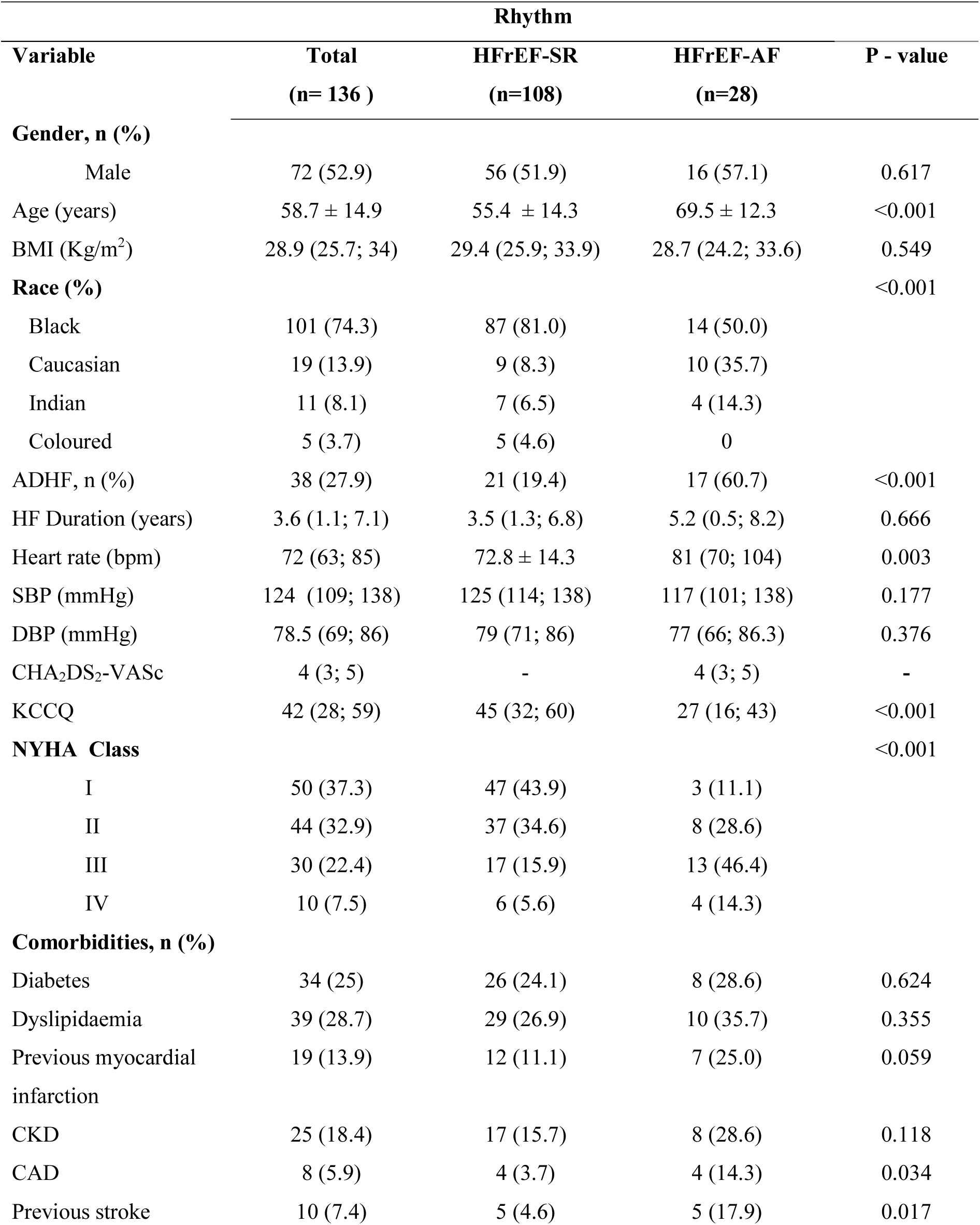
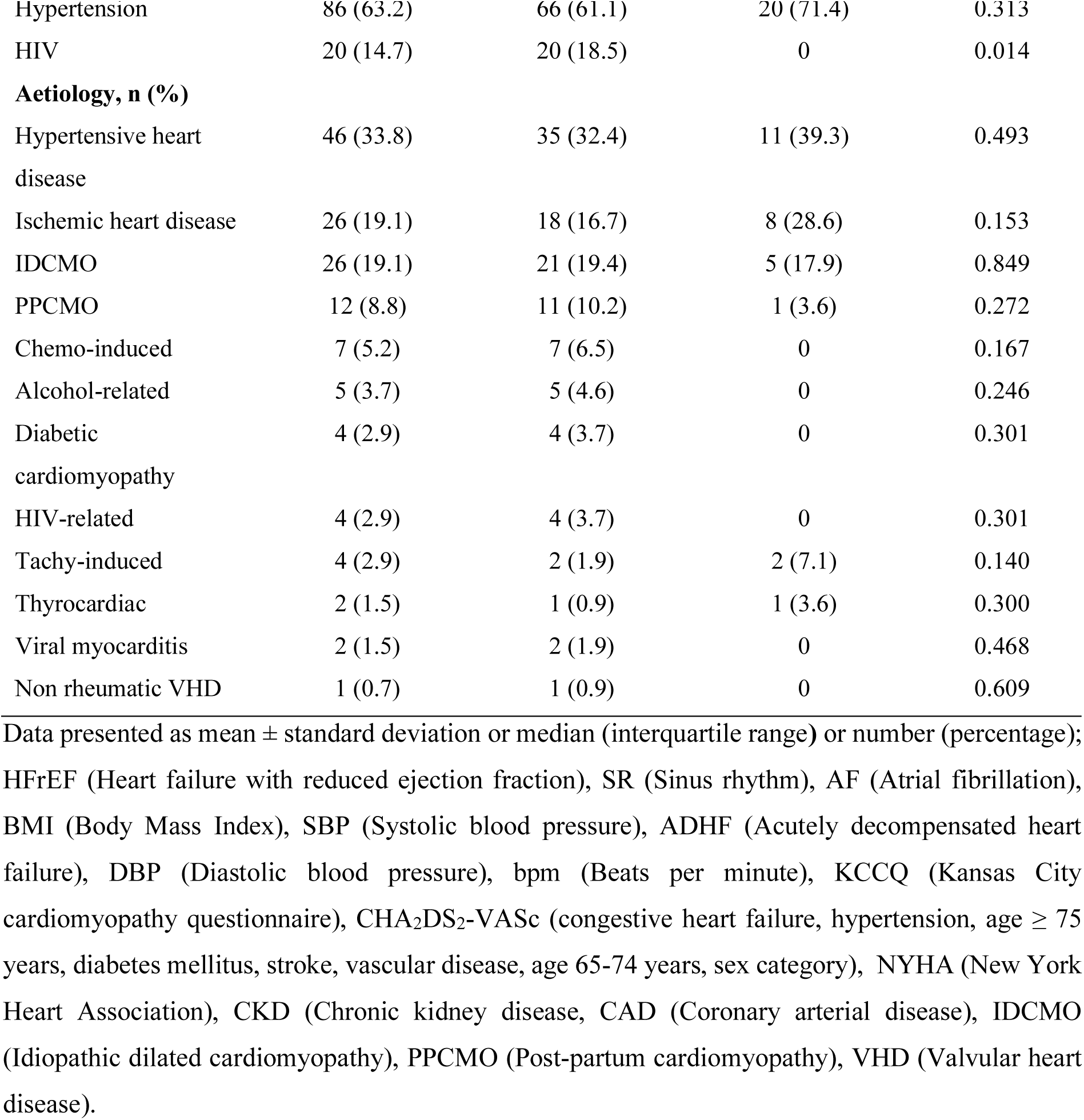
Baseline characteristics of patients with HFrEF categorised according to the presence and absence (SR) of atrial fibrillation.

The prevalence of AF in the current study was 21% (Table I). Fourteen (50%) patients with AF were of Black African ethnicity (p = 0.001). Patients with AF were significantly older (69.5 ± 12.3 years) than those in SR (55.4 ± 14.3 years; p< 0.001). Patients with HFrEF-AF had significantly lower KCCQ (27 [16-43]) compared to those in SR (45 [32-60]) (p < 0.001) as well as worsening functional class (NYHA III-IV; 60.7% vs 21.5%, p < 0.001).

Amongst patients with AF, a greater proportion had coronary artery disease (14.3 % vs 3.7%, p = 0.034) and a history of previous stroke compared to patients in SR (Table I).

### Pharmacotherapy profile of the enrolled patients with HFrEF

Beta-blockers were the most prescribed class of drugs, with 111 out of 125 patients (88.8%) receiving therapy. The prescription rate was significantly higher in patients with SR compared to those with AF (91.9% vs. 76.9%, p = 0.031). Among β-blocker users, carvedilol was the predominant agent, used in 97.3% of patients, with universal use in the AF group (100%) and 96.7% in the SR group. The median carvedilol dose was significantly lower among patients with AF compared to SR patients (15.6 mg [IQR 12.5–25] vs. 25 mg [IQR 12.5–50]; p = 0.002) (Table II).

**Table II.** Pharmacotherapy profile of enrolled patients with HFrEF stratified by heart rhythm: Sinus rhythm (SR) vs atrial fibrillation (AF).

| RHYTHM |  |  |  |  |
| --- | --- | --- | --- | --- |
| Variable | Total | HFrEF-SR | HFrEF-AF | P - value |
|  | (n= 136 ) | (n=108) | (n=28) |  |
| Medication |  |  |  |  |
| Enalapril, n (%)<br>(n=122) | 85 (69.7) | 72 (75) | 13 (50) | 0.033 |
| Enalapril dose (mg) | 10 (10; 20) | 20 (10; 20) | 10 (5; 10) | 0.004 |
| Beta-blocker, n (%)<br>(n=125) | 111 (88.8) | 91 (91.9) | 20 (76.9) | 0.031 |
| Carvedilol, n (%) | 107 (97.3) | 87 (96.7) | 20 (100) |  |
| Atenolol, n (%) | 2 (1.82) | 2 (2.22) | 0 |  |
| Bisoprolol, n (%) | 1 (0.91) | 1 (1.11) | 0 |  |
| Carvedilol dose (mg) | 25 (12.5; 50) | 25 (12.5; 50) | 15.6 (12.5; 25) | 0.002 |
| Losartan, n (%)<br>(n=124) | 18 (14.5) | 15 (15.3) | 3 (11.5) | 0.628 |
| Furosemide, n (%)<br>(n=121) | 97 (80.2) | 78 (81.3) | 19 (76) | 0.578 |
| Furosemide dose | 40 (40; 80) | 40 (40; 80) | 80 (40; 80) | 0.077 |
| Spironolactone, n<br>(%) | 86 (69.9) | 73 (75.3) | 13 (50) | 0.013 |
| Spironolactone dose | 25 (25;25) | 25 (25; 25) | 25 (25; 25) |  |
| Anticoagulation, n<br>(%) (n=119) | 24 (20.2) | 9 (9.5) | 15 (60) | <0.001 |

Data presented as mean ± standard deviation or median (interquartile range) or number (percentage); HFrEF (Heart failure with reduced ejection fraction), SR (Sinus rhythm), AF (Atrial fibrillation)

Furosemide was prescribed to 80.2% of patients, with comparable usage between rhythm groups (SR = 81.3% vs. AF = 76%; p = 0.578). Although not statistically significant, the median furosemide dose trended to be higher in the AF group (80 mg [IQR 40–80]) than in the SR group (40 mg [IQR 40–80]; p = 0.077) (Table II).

Spironolactone use differed significantly between rhythm groups, with higher usage in the SR group (75.3% vs. 50%; p = 0.013).

Anticoagulation was significantly more common in AF patients, with 60% receiving anticoagulation versus only 9.5% in the SR group (p < 0.001), reflecting appropriate management.

Enalapril was prescribed to 69.7% of patients, with significantly higher usage among SR patients (75%) compared to AF patients (50%; p = 0.033). Median enalapril dose was also higher in the SR group (20 mg [IQR 10–20] vs. 10 mg [IQR 5–10]; p = 0.004).

### Biochemistry

Estimated glomerular filtration rate (mL/min) was significantly lower in HFrEF-AF patients (52 [37-70.9]) compared with SR counterparts (71.9 [50-98.1]) (p=0.046). Baseline NT-proBNP, cTnT and creatinine levels were numerically higher in patients with HFrEF-AF but not significantly different. Baseline urea was significantly higher in patients with AF compared to those in SR (Table III).

**Table III.** Baseline biochemical parameters of patients with HFrEF categorised according to the presence and absence (SR) of AF.

| Variable | Rhythm |  |  | P - value |
| --- | --- | --- | --- | --- |
|  | Total (N=136) | HFrEF-SR<br>(n=93) | HFrEF-AF<br>(n=23) |  |
| Haemoglobin (g/dl) * | 13 ± 2.3 | 13 ± 2.2 | 13 ± 2.5 | 0.960 |
| Platelet count † (x10<sup>9</sup>/L) | 245 (195; 315) | 251 (201; 306) | 241 (193.5; 327.5) | 0.933 |
| Creatinine (µmol/l) ‡ | 99.5 (79; 137) | 97 (76; 131) | 116.5 (97; 146) | 0.060 |
| Urea (mg/dL) ‡ | 6.6 (4.6; 12.1) | 6 (4; 11.8) | 10.5 (6.1; 13.7) | 0.020 |
| eGFR (mL/min) ‡ | 65.6 (48; 95.9) | 71.9 (50; 98.1) | 52 (37; 70.9) | 0.046 |
| Total cholesterol<br>(mmol/L) § | 3.5 ± 1.1 | 3.8 ± 1.1 | 3.1 ± 1.0 | 0.058 |
| LDL (mmol/L) \|\| | 2.3 ± 0.9 | 2.4 ± 0.9 | 2.1 ± 0.8 | 0.235 |
| HDL (mmol/L) \|\| | 0.96 (0.8; 1.2) | 0.96 (0.82; 1.3) | 0.9 ± 0.2 | 0.834 |
| HBA1C (%)# | 6.9 (6.1; 7.4) | 6.5 (5.8; 7.4) | 6.9 (6.5; 7.5) | 0.447 |
| INR (mmol/L) ** | 1.27 (1.1; 1.5) | 1.16 (1.0; 1.4) | 1.4 (1.2; 2.1) | 0.040 |
| NT-proBNP (pg/mL) | 1030 (678; 1841) | 993.5 (582; 1779) | 1243 (847; 2040) | 0.164 |
| cTnT (pg/mL) | 26.5 (19.1; 36) | 26.7 (19.3; 35.3) | 28.4 (17.9; 42.9) | 0.644 |

Data presented as mean ± standard deviation or median (interquartile range); HFrEF (Heart failure with reduced ejection fraction), SR (Sinus rhythm), AF (Atrial fibrillation), eGFR (Estimated glomerular filtration rate), LDL (Low-density lipoprotein), HDL (High-density lipoprotein), HBA1C (Glycated haemoglobin), INR (International normalized ratio), NT-ProBNP (N-terminal pro brain natriuretic peptide), cTnT (Cardiac troponin T).
\*Data available for 91 patients.
† Data available for 85 patients.
‡ Data available for 96 patients.
§ Data available for 47 patients.
|| Data available for 45 patients

### Echocardiography

Patients with HFrEF-AF had a significantly greater LA volume (ml) (83.9 [64.5-96.9]) than those with HFrEF-SR (58.9 ± 25.1) (p< 0.001) (Table IV). Left atrial strain reservoir function (%) was significantly lower in patients with HFrEF-AF (29.00 ± 13.60) compared to those with HFrEF-SR (41.56 ± 15.46) (p < 0.001) (Table IV).

**Table IV.**
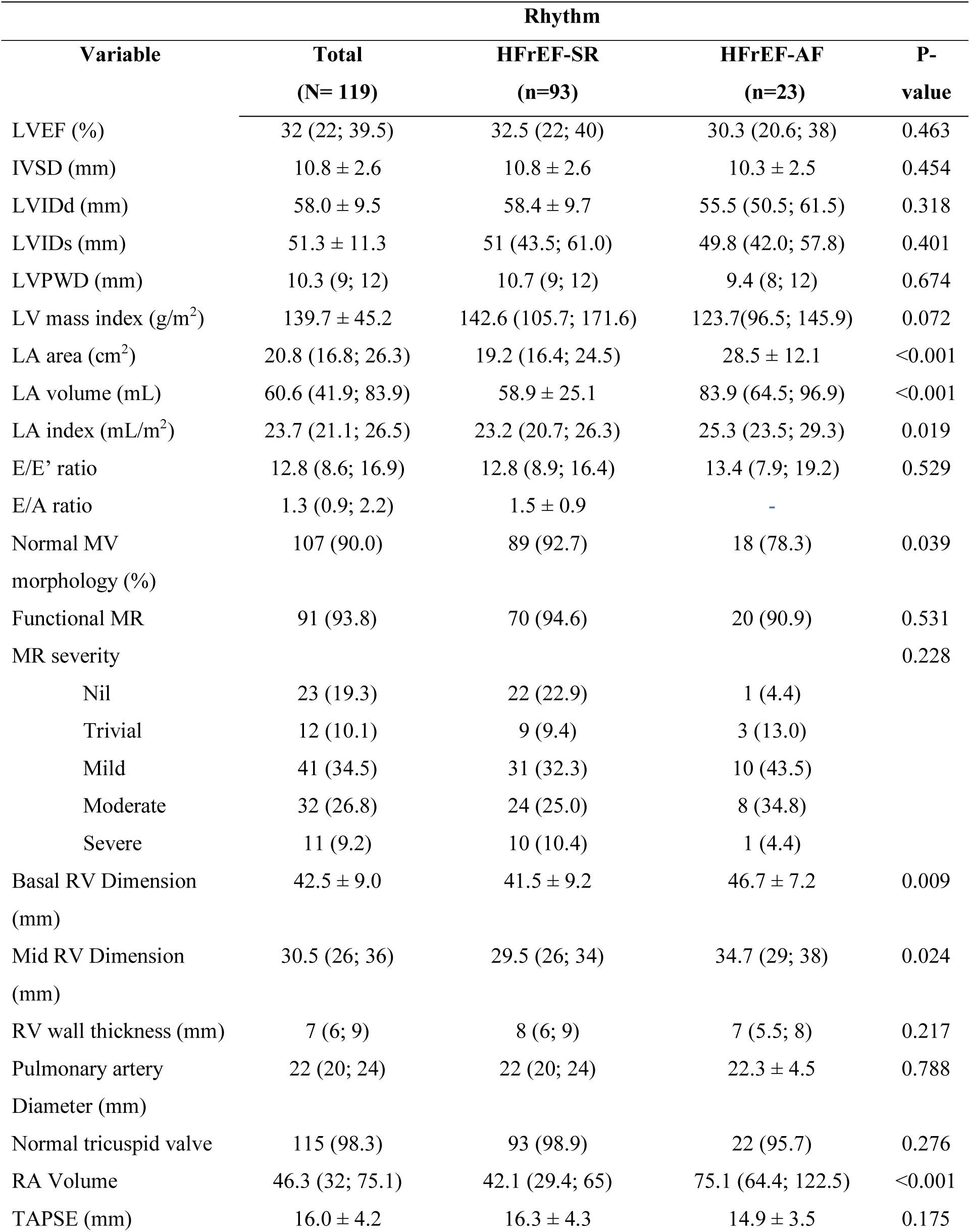
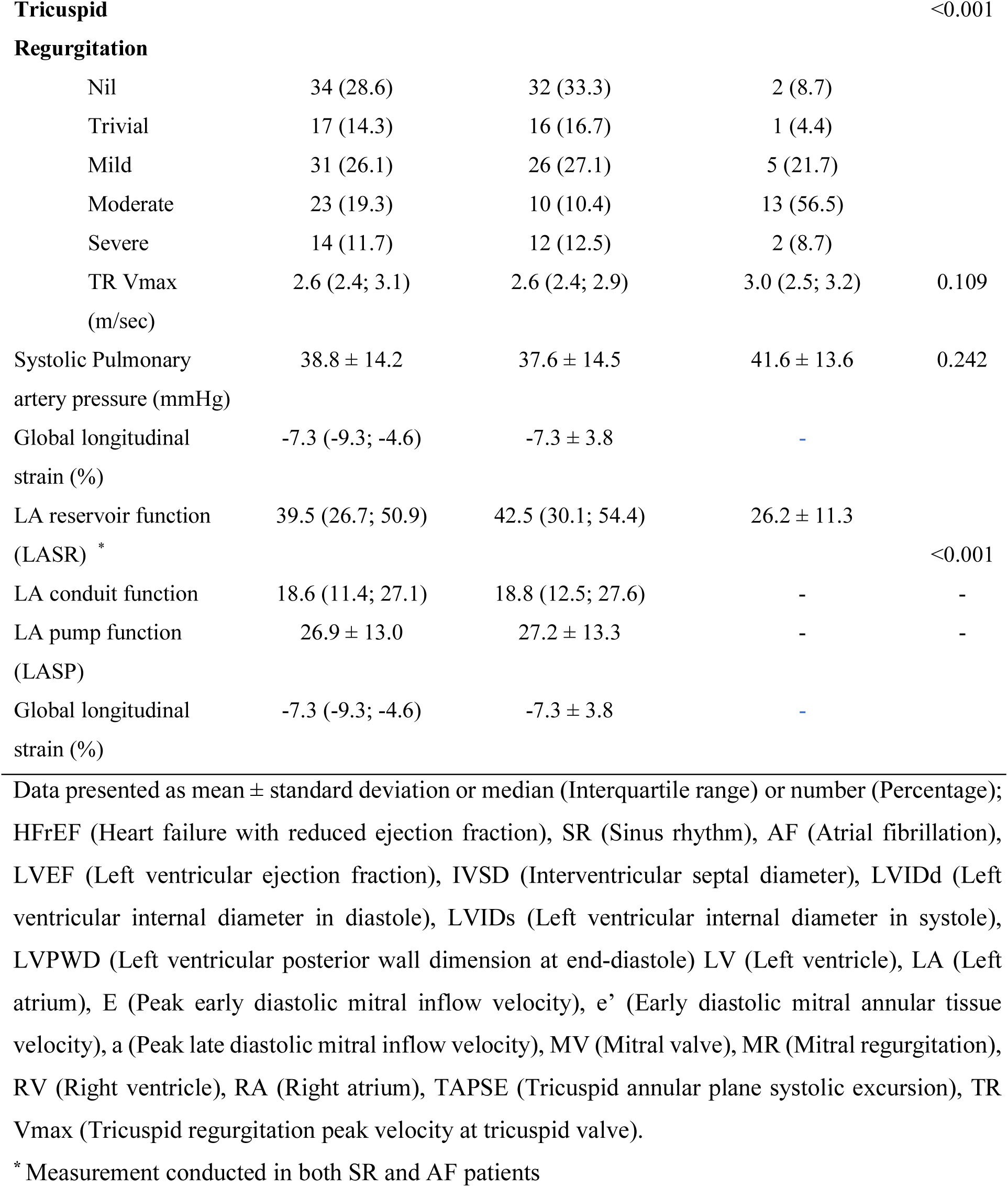
Echocardiographic parameters categorised according to the presence (SR) and absence of AF.

HFrEF-AF patients had significant right ventricular (RV) remodelling (basal RV dimension: 46.7 ± 7.2; mid RV dimension: 34.7 [29; 38]) compared to those without AF (basal RV dimension: 41.5 ± 9.2; mid RV dimension: 29.5 [26; 34]). Patients in the HFrEF- AF group had significantly greater right atrial volumes compared to the SR group (75.1 (64.4; 122.5) vs 42.1 (29.4; 65), p < 0.001). The systolic pulmonary artery pressures did not differ significantly between the two groups (Table IV).

### Mortality Outcomes

The overall study time at risk was 42 months, and the median was 30.63 months. A total of 37 patients died during the study period. Among patients who died, 40% had concomitant AF (p=0.001) (supplementary Table S1). The overall probability of survival at the end of the study was 0.66 (0.56-0.75). The probability of survival at 42 months in SR and AF was 0.73 (0.61-0.82) and 0.41 (0.22-0.61) (p<0.000), log-rank) respectively. The median survival time for patients in AF was 27.1 months. The majority of patients in SR survived beyond the median survival time at risk (Figures 2).

**Figure 2.**
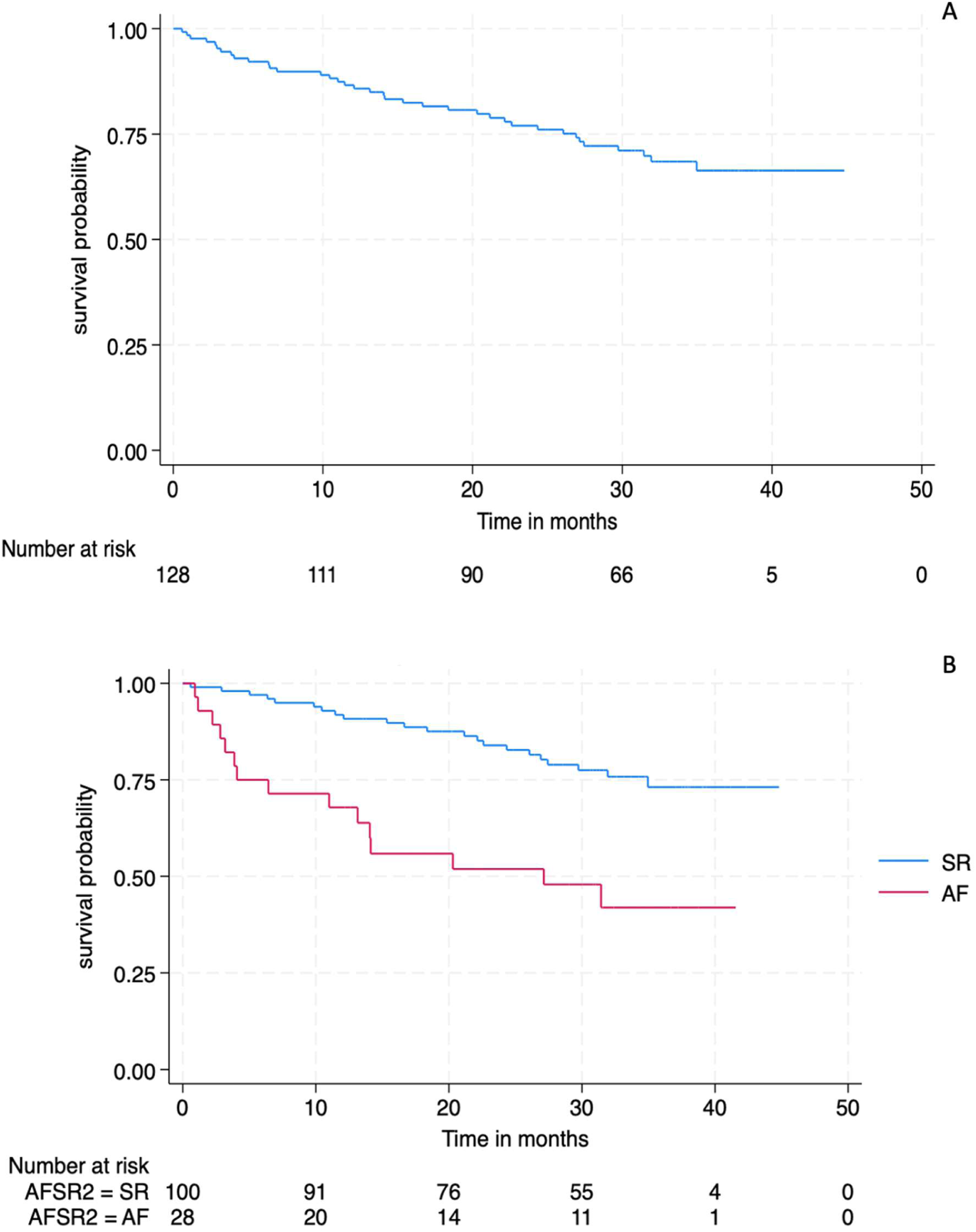
A Kaplan Meier plots illustrating the survival rate of patients enrolled in the study. Panel A illustrates the overall survival rate of patients. Panel B, illustrates the survival rate of patients with atrial fibrillation (red) compared to those in sinus rhythm (blue) (p<0.001, log rank).

The survival rate of patients with HFrEF-SR at 12 months was 0.92 (CI: 0.84-0.96) and that of HFrEF-AF patients was 0.68 (CI:0.47-0.82) (Table V).

**Table V.** Survival rate of patients over 3-month intervals within the first year of survival time at risk.

| **Survival time at Risk (Months)** | ***Survival Rate (95%CI)*** |
| --- | --- |
| **HFrEF-SR** |  |
| 3 | 0.98 (0.92-0.99) |
| 6 | 0.97 (0.91-0.99) |
| 9 | 0.95 (0.88-0.98) |
| 12 | 0.92 (0.84-0.96) |
| 15 | 0.91(0.83-0.95) |
| **HFrEF-AF** |  |
| 3 | 0.86 (0.66-0.94) |
| 6 | 0.75 (0.55-0.87) |
| 9 | 0.71 (0.51-0.85) |
| 12 | 0.68 (0.47-0.82) |
| 15 | 0.57 (0.37-0.73) |

HFrEF (Heart failure with reduced ejection fraction), SR (Sinus rhythm), AF (Atrial fibrillation), CI (Confidence interval)

In a multivariate logistic regression model (R^2^ =0.43, p < 0.001) tricuspid annular plane systolic excursion (TAPSE), heart rate, early diastolic mitral inflow velocity/ early diastolic mitral annular tissue velocity ratio (E/e’) and prior stroke were independently associated with all-cause mortality (Table VI). A higher heart rate (Odds ratio (OR) =1.05 [1.00- 1.10], p= 0.028) and E/e’ (1.24 [1.05- 1.47], p= 0.012), a measure of diastolic dysfunction, were associated with increased odds of death. A prior history of stroke was a strong predictor of death (OR= 129.49 [2.44-1503], p = 0.014). Conversely, elevated TAPSE (OR = 0.77 [0.59-0.99], p = 0.039), a surrogate marker for right ventricular function, was protective.

**Table VI.** Univariate and multivariable logistic regression analysis for predictors of all- cause mortality.

| Variable | Univariate |  | *Multivariate |  |
| --- | --- | --- | --- | --- |
|  | Odds Ratio (95% CI) | p- value | Odds Ratio (95% CI) | P- value |
| Age | 1.03(1.004-1.06) | 0.022 |  |  |
| Heart rate | 1.04 (1.01-1.07) | 0.002 | 1.05 (1.00- 1.10) | 0.028 |
| KCCQ | 0.94 (0.92-0.97) | <0.001 |  |  |
| ADHF | 4.5 (1.94-10.4) | <0.001 |  |  |
| Diabetes | 2.1 (0.89-4.8) | 0.088 |  |  |
| Previous stroke | 4.3 (1.1-16.1) | 0.033 | 129.5 (2.44-1503) | 0.014 |
| NT-PROBNP | 1.00 (1.00-1.00) | 0.031 |  |  |
| eGFR | 0.97 (0.96-0.99) | 0.004 |  |  |
| Creatinine | 1.01 (1.00-1.02) | 0.015 |  |  |
| Chronic kidney disease | 3.2 (1.3-8.0) | 0.013 |  |  |
| LVEF | 0.93 (0.89-0.98) | 0.002 |  |  |
| LVIDS | 1.04 (1.01-1.09) | 0.025 |  |  |
| E/e' | 1.1 (1.02-1.2) | 0.018 | 1.24 (1.05- 1.47) | 0.012 |
| Basal RV dimension | 1.1 (1.0-1.1) | 0.003 |  |  |
| TAPSE | 0.79 (0.7-0.9) | 0.001 | 0.77 (0.59- 99) | 0.039 |
| RA volume | 1.01 (1.00-1.02) | 0.006 |  |  |
| Systolic pulmonary artery pressure | 1.03 (0.99-1.1) | 0.127 |  |  |
| LA reservoir strain | 0.93 (0.91-0.97) | 0.001 |  |  |

BMI (Body mass index), KCCQ (Kansas City Cardiomyopathy Questionnaire), MR (Mitral regurgitation), ADHF (Acute decompensated heart failure), NT-proBNP (N-terminal pro natriuretic peptide), eGFR (Estimated glomerular filtration rate), LVEF (Left ventricular ejection fraction), LVIDs (Left ventricular internal diameter in systole), LA (Left atrium), E (Early diastolic mitral inflow velocity), e' (Early diastolic mitral annular tissue velocity), RV (Right ventricle), TAPSE (Tricuspid annular plane systolic excursion), RA (Right atrium), GLS (Global longitudinal strain).
\*Backwards elimination using a p-value = 0.15

In an age-adjusted regression model, KCCQ score, heart rate, E/e’, and prior stroke were significantly associated with all-cause mortality (R² = 0.40, p < 0.001). A higher quality of in the two weeks preceding presentation (as indicated by a lower KCCQ score) was protective (OR = 0.94 [0.90–0.99], p = 0.026). TAPSE showed a trend toward being protective but did not reach statistical significance (p = 0.070) (Table VII).

**Table VII.** Age-adjusted multivariate logistic regression model of variables associated with all-cause mortality.

| Variables | Univariate |  | Multivariate |  |
| --- | --- | --- | --- | --- |
|  | Odds ratio (95% CI) | p-value | Odds ratio (95% CI) | P-value |
| Age | 1.03 (1.0-1.06) | 0.022 | 0.98 (0.92- 1.04) | 0.965 |
| TAPSE | 0.79 (0.7-0.9) | 0.001 | 0.80 (0.64- 1.02) | 0.070 |
| Heart rate | 1.04 (1.0-1.1) | 0.002 | 1.04 (1.00- 1.09) | 0.047 |
| E/e' | 1.1 (1.02-1.2) | 0.018 | 1.28 (1.05- 1.54) | 0.014 |
| Previous stroke | 4.3 (1.1-16.1) | 0.033 | 443 (3.07- 641) | 0.016 |
| KCCQ | 0.94 (0.92-0.97) | <0.001 | 0.94 (0.89- 0.99) | 0.026 |

CI (Confidence interval), TAPSE (Tricuspid annular plane systolic excursion), E (Early diastolic mitral inflow velocity), e' (Early diastolic mitral annular tissue velocity), KCCQ (Kansas City Cardiomyopathy Questionnaire).

## Discussion

This study compared the clinical characteristics of South African patients with HFrEF-SR and HFrEF-AF as well as the factors associated with their mortality over a 30 month period. Our findings demonstrate that (1) Patients with HFrEF-AF were significantly older than patients in SR; (2) The estimated prevalence of HFrEF-AF was comparatively low (21%) in this cohort; (3) Patients with HFrEF-AF had advanced structural heart disease compared to those in SR, (4) Patients with HFrEF-AF, in this cohort, received less intensive pharmacotherapy administered at lower doses compared to their SR counterparts (5). Patients with HFrEF-AF had higher morbidity and mortality compared to their SR counterparts.

The mean age of HFrEF patients was 58.7 ± 14.9 years, which is comparable to findings in other SSA countries (20, 21). Patients with HFrEF-AF were significantly older than those in SR by over a decade, consistent with global trends (22). Our cohort of HFrEF-AF patients was significantly younger than their North American and European counterparts, despite a similar trend towards advanced age (22–24). We postulate that this is largely attributable to the high prevalence of a more malignant form of long-standing hypertension in Black South Africans (25). Hypertension is an established etiology of HFrEF. Given the well-documented genetic predisposition to salt sensitivity in individuals of Black African ethnicity as well as increased dietary salt intake, hypertension manifests at a younger age and presents with greater severity (26, 27). Furthermore, patients in this demographic are more likely to present with hypertension-mediated organ damage compared to other racial groups (25, 28). An overactive Renin-Angiotensin-Aldosterone System (RAAS) and sympathetic nervous system are pathophysiological hallmarks of salt sensitivity as well as HFrEF, potentially exacerbating disease progression (29, 30). This is further compounded by poor blood pressure control and limited access to healthcare in our setting (7, 31, 32). Approximately 15.4% of individuals achieve adequate blood pressure control in South Africa, a figure significantly lower compared to other regions globally (7, 31, 32). A recent study demonstrated that the index presentation of hypertension-mediated organ damage in SSA is approximately 15 years younger than in patients in HICs (5, 21). Thus, HFrEF is often a manifestation of severe, long-standing, poorly controlled hypertension.

The prevalence of HFrEF-AF in this study was comparable to other low- to middle-income countries (12-20%) but markedly lower than that of HICs (20, 22). Our findings suggest that the burden of HFrEF-AF, in South Africa, is a function of advanced, severe HF rather than advancing age. The prevalence of HFrEF-AF, in HICs, is largely driven by advanced age (33). This is supported by a longer average life expectancy of approximately 80 years in HICs compared to approximately 60 years in South Africa, as well as a larger geriatric population compared to SSA (34). In addition to this, we postulate that a large proportion of our patients may succumb to mortality before reaching a tertiary institution or, although known to our facility, may be critically ill and demise outside of our institution, resulting in loss to follow-up. The significantly lower KCCQ score and low survival rate within the first 12 months observed in patients with HFrEF-AF in this study lend support to this hypothesis. The ARIC study showed that African Americans had a 41% lower age-adjusted and gender-adjusted risk of AF compared with Caucasians, despite a greater risk factor burden amongst African Americans (35). A similar racial variation was observed in the United Kingdom (36). This may also be a contributing factor to the low AF prevalence observed in our cohort. These observations, highlight the potential complexity of the underlying pathophysiological mechanisms of AF in the setting of HFrEF. Exploring these potential pathophysiological undertones may provide valuable insights for managing this unique demographic.

The presence of AF in HFrEF was associated with a more advanced form of HFrEF as evidenced by significantly lower KCCQ scores, higher functional class (NYHA), worser echocardiographic indices of disease severity such as LA volume, strain and extensive right ventricular remodelling as well as numerically higher levels of NT-proBNP in comparison to those in SR. Our findings suggest that these patients with HFrEF-AF have extensive structural heart disease, severe symptomatology and are more likely to deteriorate rapidly. A similar impact of AF in HFrEF with respect to quality of life and mortality is observed globally (33, 37, 38). The onset of AF in HFrEF is facilitated by adverse structural and electrophysiological remodelling of the atria. Furthermore, AF exacerbates these pathophysiological processes, via increased ventricular heart rate, loss of atrial systolic function and subsequent decrease in cardiac output eliciting an increase in HF symptoms, worsening cardiac function, morbidity and mortality (33, 39).

Despite representing a subgroup with greater morbidity and severity, patients with HFrEF-AF received suboptimal guideline-directed medical therapy (GDMT) compared to their SR counterparts. Beta -blockers were the most frequently prescribed agents overall but their use was significantly lower in HFrEF-AF patients compared to those in SR, and the median carvedilol dose substantially lower in the AF group. A similar trend was observed for angiotensin converting enzyme inhibitors (ACE-I) and spironolactone. This is clinically important, as GDMT including β -blockers, has been shown to reduce mortality and improve functional status in HFrEF, regardless of rhythm status (8). Suboptimal GDMT titration in HFrEF-AF may reflect clinical inertia in complex multimorbid patients, hesitancy to initiate or up-titrate GDMT in the context of acutely decompensated patients due to hemodynamic instability, renal dysfunction or fear of low tolerance amongst AF patients (40). Furthermore, the significantly higher use of oral anticoagulation (OAC) in HFrEF- AF patients reflects appropriate stroke prevention measures. However, the finding that only 60% of HFrEF-AF patients were anticoagulated, despite clear indication, suggests barriers to implementation of OAC or possible contraindications. Factors may include concerns regarding bleeding risk, poor INR control with warfarin (the only available OAC in the public sector at the time of enrolment), adherence challenges, limited access to regular monitoring, and healthcare provider caution (41). These pharmacological disparities, may partially explain the excess morbidity and mortality observed in the HFrEF-AF patient group and highlight potential opportunities to strengthen inpatient and early outpatient optimisation of GDMT.

The finding that Individuals with HFrEF-AF had a significantly reduced survival rate compared to patients in SR supports the hypothesis that HFrEF-AF present with more severe clinical manifestations. In this cohort, a large percentage of patients with HFrEF-AF presented in acute decompensated HF, and succumbed to the outcome of all-cause mortality within the first year. The evidence regarding whether AF itself is an independent predictor of all-cause mortality in HFrEF is inconsistent. A large European multinational long-term registry reported that AF was not independently associated with all-cause mortality (33). It is worth noting however, that given our study design, we could not determine this.

In light of the extent of structural heart damage as well as clinical severity in patients with HFrEF-AF, other variables better performed in predicting the outcome of all-cause mortality. Worsening diastolic dysfunction, increasing heart rate, prior stroke and lower quality of life two weeks prior to presentation were independently associated with an increased risk of all-cause mortality, while better right ventricular function was protective. These findings support the view that in the setting of HFrEF, AF serves as a marker of disease severity rather than as an independent predictor of mortality. There are few studies in SSA that have been able to establish AF as an independent predictor of all-cause mortality (42, 43). While these studies had a much larger sample size, their populations were predominantly heterogeneous HF (Malamba et al) and AF populations (Mandi et al.) (42, 43). Furthermore, there is now recent evidence to suggest a much more significant burden of AF in heart failure with preserved ejection fraction (33, 44).

### Clinical Perspective and Future directions

To the best of our knowledge, this study is the first prospective observational study to report on the clinical impact of AF on HFrEF in SSA patients. Despite the lower prevalence of HFrEF-AF (relative to HICs), associated with a younger age of onset and has increased morbidity and mortality in this demographic. These observations have important clinical and public health implications, particularly in resource-limited settings where hypertension and late presentation remain major contributors to the burden of HFrEF.

Additional large-scale, racially diverse multi-centre prospective studies are required to accurately define the clinical impact of AF in HFrEF within this high-risk population. Studies should determine whether the lower observed prevalence of HFrEF-AF is due to underdiagnosis, differences in access to healthcare, or population-specific characteristics. The gaps in pharmacotherapy identified, underscore a need for early GDMT optimisation as well as rhythm-specific strategies with a structured follow-up in outpatient settings. Further studies investigating the impact of GDMT, catheter ablation, and anticoagulation strategies in this unique subpopulation could provide insights into improving clinical outcomes. Should these findings be replicated in subsequent studies, they may support the necessity for earlier and more aggressive interventions in this demographic of patients.

### Limitations

Our study had several limitations. The main limitation of the study was the small sample size. Recruitment was disrupted due to the COVID-19 pandemic lockdown regulations. In AF detection, we used standard 12-lead ECG and did not conduct prolonged cardiac rhythm monitoring using Holter monitors or loop recorders. Prolonged monitoring would have enabled the detection of paroxysmal AF. This was a cross-sectional study with prospective follow-up; most of the AF in our study was permanent. The study is a single-centre study done at a tertiary institution, and hence, our findings may not be generalisable to the general population.

## Conclusion

Our study has demonstrated that AF is common in patients with HFrEF but comparatively low relative to HICs, and is associated with a more severe clinical presentation and a high mortality rate in comparison to their SR counterparts.

## Data Availability

The data that support the findings of this study are available from the corresponding author upon reasonable request. Access to data may be subject to approval by the institutional ethics committee and compliance with applicable data protection and confidentiality requirements.

## Supplementary Material

Table S1

